# Beyond Binary Vasospasm: A Continuum Model Relating Severity and Distribution to Perfusion Deficits After Aneurysmal SAH

**DOI:** 10.64898/2026.07.16.26358285

**Authors:** Christian Thaler, Bogdana Tokareva, Lukas Meyer, Christian Heitkamp, Helge Kniep, Vincent Geest, Lasse Dührsen, Hanno S. Meyer, Maxim Bester, Jens Fiehler, Felix Schlicht

**Author notes:** Corresponding author University Medical Center Hamburg-Eppendorf Martinistrasse 52, 20246 Hamburg-Eppendorf Germany.

## Abstract

**Background:** Cerebral vasospasm is a frequent complication after aneurysmal subarachnoid hemorrhage (aSAH) and is associated with delayed cerebral ischemia (DCI) and unfavorable outcome. While CTA-based vasospasm grading is frequently used, its relationship with actual cerebral perfusion remains incompletely understood. This study investigates the association between vasospasm severity and distribution and territorial perfusion deficits.

**Methods:** In this retrospective single-center study, 513 CT examinations (CTA and CT perfusion) from 194 patients with aSAH were analyzed. Vasospasm was graded per vessel segment using the CTA Vasospasm Score, and perfusion deficits were assigned to corresponding vascular territories (left/right anterior circulation, posterior circulation). Vasospasm distribution was further classified by severity and multifocality. Associations between vasospasm score and perfusion deficits were assessed using a generalized linear mixed model with binomial distribution, adjusting for Hunt & Hess grade, modified Fisher score, and days since hemorrhage.

**Results:** Vasospasm was detected in 79.3% of examinations, and a perfusion deficit in at least one territory was present in 62.6%. The proportion of perfusion deficits increased progressively with both vasospasm severity and multifocality, ranging from 21.7-25.0% in the absence of vasospasm to 81.2-82.2% in severe multifocal vasospasm. The CTA Vasospasm Score was significantly associated with perfusion deficits in all territories (OR 1.36-1.50), with stronger associations in the anterior than posterior circulation.

**Conclusion:** Vasospasm severity and distribution are strongly associated with perfusion deficits, supporting a continuum model of ischemic risk. However, the substantial proportion of perfusion deficits occurring independent of vasospasm suggests additional microcirculatory mechanisms not captured by CTA. CT perfusion should be considered complementary to CTA, particularly in clinically deteriorating or non-assessable patients.

## Introduction

Cerebral vasospasm remains a frequent complication after aneurysmal subarachnoid hemorrhage (aSAH) and is associated with delayed cerebral ischemia (DCI) and worse outcomes.[1, 2] Besides neurological examinations, current surveillance strategies are based on transcranial Doppler (TCD) as well as vascular imaging with CT angiography (CTA) or digital subtraction angiography (DSA), as recommended by the American Heart Association and American Stroke Association.[3] These diagnostic tools visualize the degree of arterial narrowing, i.e. grade of vasospasm, and can help to guide further treatment decision, such as endovascular drug administration or even PTA in selected cases, to prevent cerebral infarction and unfavorable functional outcomes.[4] In an attempt to quantify and classify vasospasm in a more standardized way the CTA Vasospasm Score has been introduced by van der Harst et al. by assessing the degree of vasospasm of 17 vessel segments, scored as 0 for no visible vasospasm, 1 for mild (<50% arterial narrowing) and 2 for severe (>50% arterial narrowing) vasospasm. Hereby, the score helped to identify patients with a high risk of developing DCI and unfavorable outcomes.[5]

However, the pathophysiological mechanisms behind DCI are multifactorial and besides arterial narrowing, factors such as cerebral perfusion pressure / altered autoregulation, collateral status, microcirculatory dysfunction, microthrombosis, neuroinflammation, and cortical spreading depolarizations may play a role in the development of DCI. CTA does not provide direct insight into perfusion status or microcirculatory changes of the brain tissue.[6] Therefore, the recent European Stroke Organization guidelines suggest to perform head CT in combination with CT perfusion (CTP), rather than CTA.[7] CTP provides further information about macro- and microvascular blood flow and provides greater diagnostic value than vessel imaging alone for identifying patients at risk of DCI.[6]

Although cerebral vasospasm is generally associated with perfusion deficits in the affected vascular territories, discrepancies are frequently observed. Some patients demonstrate perfusion deficits despite the absence of vasospasm on CTA, whereas others exhibit no perfusion abnormalities despite significant vasospasm.[8] The aim of this study is to further elucidate the relationship between cerebral vasospasm and perfusion anomalies in patients after aSAH. We hypothesized that the probability of a perfusion deficit rises progressively with both the severity and the multifocality of vasospasm.

## Patients and Methods

This retrospective, single-center study was approved by the local ethics committee (Ärztekammer Hamburg, Germany) and conducted in line with the Declaration of Helsinki. Patient consent was waived due to the retrospective nature of the study. All data were deidentified and anonymized.

### Study population

We included all adult patients (age > 18 years) treated at our tertiary stroke center for aneurysmal subarachnoid hemorrhage (aSAH) between January 2019 and December 2024. Inclusion criteria were: (1) acute SAH due to a ruptured intracranial aneurysm, (2) at least one CTP examination during hospitalization. Patients with SAH from other causes or without evidence of a ruptured aneurysm were excluded. Also, examinations with insufficient image quality were excluded. 348 patients were admitted to our hospital with aSAH during this period. Of these, 194 patients underwent at least one CT examination, including CTA and CTP, during hospitalization. Five imaging studies were excluded due to insufficient image quality. Overall, 513 CT examinations were included and analyzed.

Baseline demographic data and clinical data were extracted from electronic medical records. Hemorrhage severity was graded using the Hunt & Hess scale, and the modified Fisher scale was used to assess the amount and distribution of subarachnoid and intraventricular blood.

### Imaging Protocol

Time points and indication for each CT scan was retrieved from the electronic medical records. All CT scans were acquired using a 256-slice dual-source scanner (Somatom Definition Flash; Siemens). Acquisition parameters are detailed in the Online Supplemental Data. For each patient, the dataset comprised non-enhanced axial cranial CT (NECT) images, thin-section axial CTA, and axial, coronal, and sagittal MIP reconstructions. The imaging parameters were as follows: NECT: 120 kV, 280-340 mA, 1 mm increment, 5.0 mm slice reconstruction; CTA: 120 kV, 260-300 mAs, 1-mm increment, 5.0-mm slice reconstruction, 80 mL highly iodinated contrast medium and 50 mL NaCl flush at 4 mL/second; CTP: 80 kV, 200-250 mA, 5 mm slice reconstruction (max. 10 mm), slice sampling rate 1.50 s (min. 1.33 s), scan time 45 s (max. 60 s), biphasic injection with 30 ml (max. 40 ml) of highly iodinated contrast medium with 350 mg iodine/ml (max. 400 mg/ml) injected with 6 ml/s followed by 30 ml sodium chloride chaser bolus.

### Vasospasm Scoring and Perfusion Analysis

We used the CTA Vasospasm Score, introduced by van der Harst, to grade the severity of cerebral vasospasm.[5] In summary, vasospasm was graded as absent, mild (<50% luminal narrowing), or severe (>50% luminal narrowing), with the admission CTA serving as the reference for assessing vasospasm on follow-up CTA. Arterial segments were predefined as the supraclinoid internal carotid artery (ICA); the M1 and M2 segments of the middle cerebral artery (MCA); the A1 and A2 segments of the anterior cerebral artery (ACA); the P1 and P2 segments of the posterior cerebral artery (PCA); the basilar artery and the V4 segments of the vertebral artery.

CTP data were processed using the CT Neuro Perfusion module on a syngo.via workstation (software version VB80F; Siemens Healthineers, Forchheim, Germany). Focal perfusion abnormalities attributable to the primary hemorrhagic event or surgical intervention were excluded from consideration as DCI-related perfusion deficits. Delayed perfusion was defined as a visually apparent regional prolongation of Tmax and time-to-drain relative to the contralateral hemisphere and was assigned to vascular territories (left/right ACA, MCA, and PCA; cerebellar territory; and anterior and posterior border zone territories).

CTA and perfusion maps were independently reviewed by two experienced readers (with 5 and 11 years of neuroradiological experience), both blinded to clinical parameters and outcomes. For vasospasm and perfusions abnormalities, consensus judgment was determined after reviewing the images independently.

### Statistics

IBM SPSS 30.0. (IBM Corp., Armonk, NY, USA) and R (Version 4.5.1; R Core Team, 2025) were used for statistical analysis. Descriptive statistics are presented as frequencies and proportions (percentages [%]) for categorical variables. The association between the CTA Vasospasm Score and the presence of a perfusion deficit was assessed using binary logistic regression. As multiple examinations were available per patient, a Generalized Linear Mixed Model (GLMM) with binomial distribution and logit link function was used, including a random intercept for patient ID to account for within-patient correlation. The model was additionally adjusted for the Hunt & Hess grade, number of days since initial hemorrhage and the modified Fisher scale as fixed effects. To evaluate whether the observed associations were robust to the timing of imaging relative to the classic vasospasm window, we performed a sensitivity analysis restricting the dataset to CT examinations performed between day 3 and day 14 after the initial hemorrhage using the same GLMM.

Vessel segments that could not be reliably evaluated due to susceptibility artifacts from aneurysm clips or coils, prior vessel sacrifice, or anatomical variants (e.g., hypoplastic or absent A1/P1 segments) were excluded from the corresponding segment-level analysis. As the generalized linear mixed model estimates parameters via maximum likelihood using all available observations, these missing segment-level data were handled under a missing-at-random assumption without the need for imputation.

## Results

We included 513 CT examinations, including CTA and CTP, from 194 patients with aSAH. Mean patient age was 55.1 (12.9) years. The median Hunt & Hess grade was 3 (IQR 2-4), and the modified Fisher score 3 (IQR 2-4). DCI was diagnosed in 129 patients. A median of 2 (IQR 1-3) CT scans were performed per patient. Imaging was performed at a median of 9 (IQR 5-13) days.

Indications for CT imaging included new neurological deficits or decreased consciousness in 251 studies (48.9%), elevated transcranial Doppler velocities in 118 (23%), and routine surveillance in comatose patients in 144 (28.1%) studies.

### CTA Vasospasm Score and Perfusion Analysis

In 407 (79.3%) examinations vasospasm (grade 1 or 2) of at least one vessel segment was detected and in 321 (62.6%) examination a perfusion deficit was detected in at least one vascular territory or border zone. The vessel segments with the highest rates of vasospasm were the M1 and A1 segment. Vasospasm was least frequently detected in the V4 segment. A complete overview is given in Table1. Correspondingly, perfusion deficits were most frequently detected in the anterior border zone (51.5%), while perfusion deficits were least frequently detected in the cerebellum (2.6%).

In 106 examinations no vasospasm was detected at all. Still, in 23 of these cases (21.7%) a perfusion deficit was detected. In 120 examinations the highest scored vasospasm grade was 1 and in 66 (55%) of these cases a perfusion deficit was detected. In 287 examinations at least one vessel segment was scored with a vasospasm grade of 2 and in 232 (80.8%) of these cases a perfusion deficit was detected (Figure 1).

**Figure 1.**
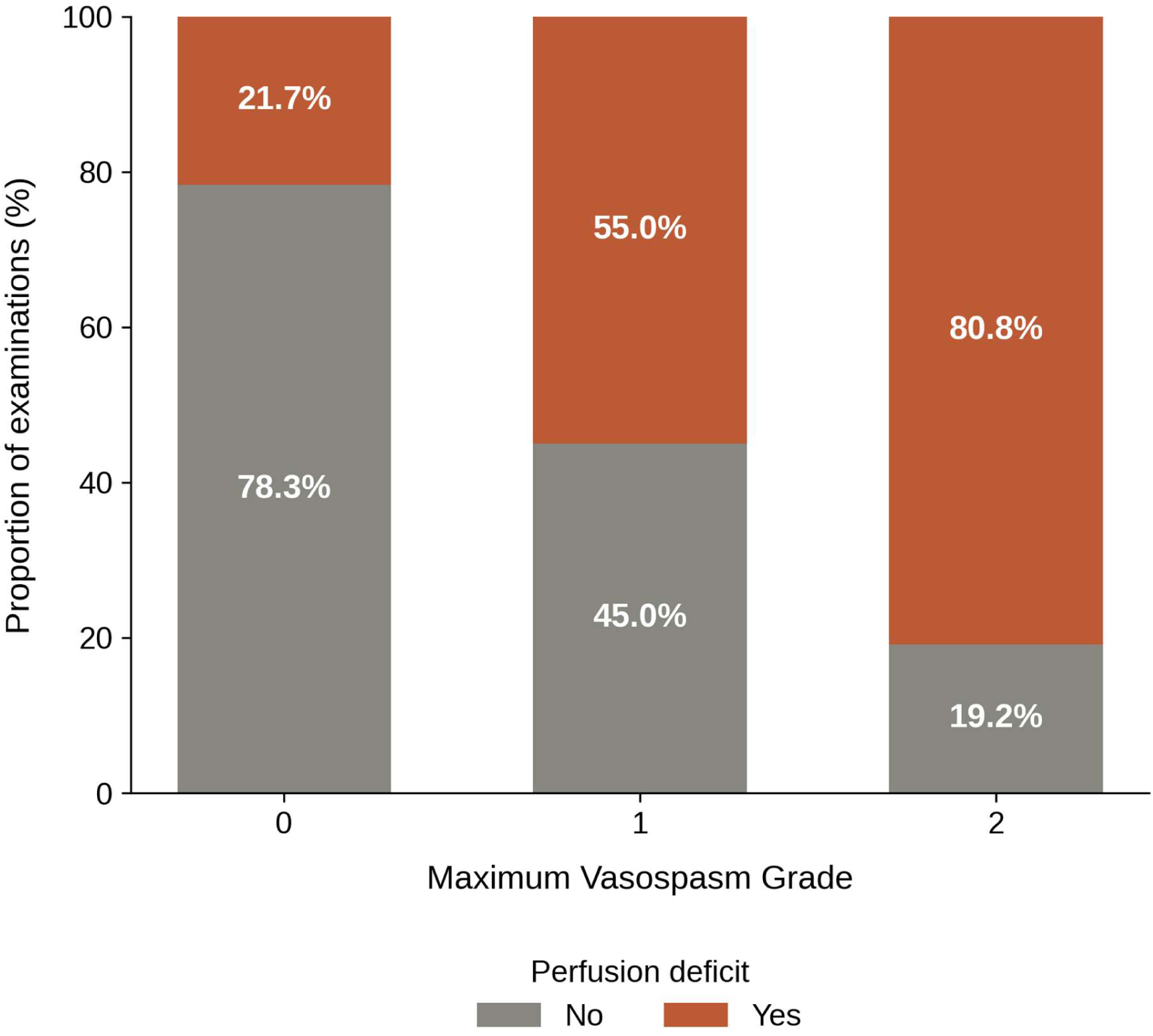
Proportion of examinations with a perfusion deficit in at least one territory as a function of the maximum detected vasospasm grade.

### Vasospasm Severity and Pattern

For further analysis we combined vessel segments and perfusion territories into three main territories: Left and right anterior circulation and posterior circulation. The vessels of the anterior circulation of one side included the ipsilateral ICA, M1 and M2 as well as A1 and A2 segment. Perfusion deficits in the MCA and ACA, as well as the anterior and posterior border zones were attributed to these vessel segments. The vessel of the posterior circulation included both V4 segments, the basilar artery as well as the P1 and P2 segments of both sides.

We classified vasospasm distribution patterns within vessel territories into 5 categories: 0 = no vasospasm, 1 = mild focal vasospasm, i.e. only one vessel segment scored with vasospasm grade 1 (<50%), 2 = mild multifocal vasospasm, i.e. ≥ two vessel segments with vasospasm grade 1, 3 = severe focal vasospasm, i.e. only one vessel segment scored with vasospasm grade 2 (>50%), 4 = severe multifocal vasospasm, i.e. ≥ two vessel segments with vasospasm grade 2. The proportion of perfusion deficits increased progressively with increasing vasospasm severity in both the right and left anterior circulation (see Figure 2). While perfusion deficits were present in only 22.4% and 25.0% of examinations without vasospasm, this proportion rose to 82.2% and 81.2%, respectively, in cases of severe multifocal vasospasm. A similar stepwise trend was observed across all intermediate categories (mild focal, mild multifocal, and severe focal vasospasm) in both territories.

**Figure 2.**
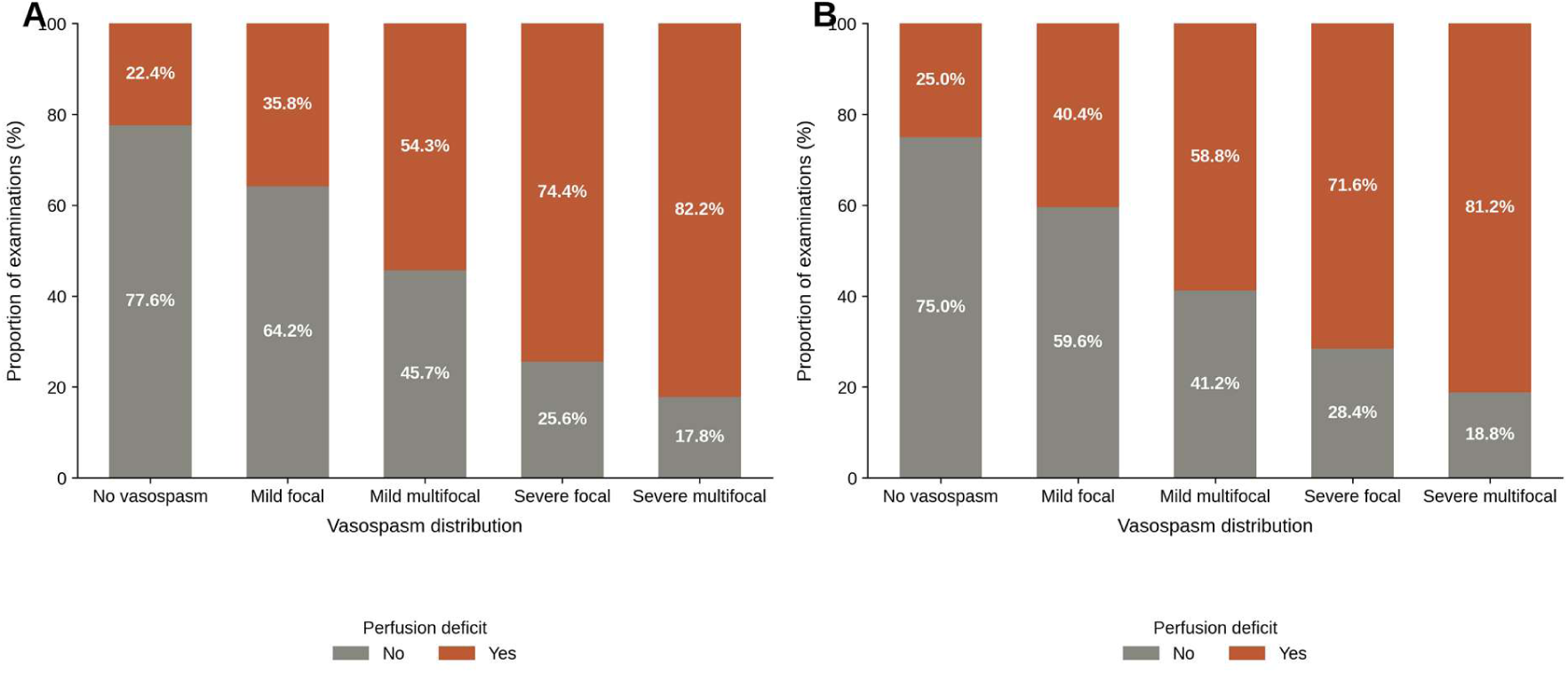
Proportion of examinations with a perfusion deficit in the left anterior circulation (A) and right anterior circulation (B) as a function of the vasospasm severity and pattern.

### CTA Vasospasm Score and Perfusion Deficits

With increasing CTA Vasospasm Score, the proportion of perfusion deficits within the territory of the affected vessel increased correspondingly. In the posterior territory, the distribution was more heterogeneous; nevertheless, a similar trend emerged, with patients with higher CTA Vasospasm Scores more frequently showing perfusion deficits. Figure 3 shows the rates of perfusion deficits in relation to the CTA Vasospasm Score.

**Figure 3.**
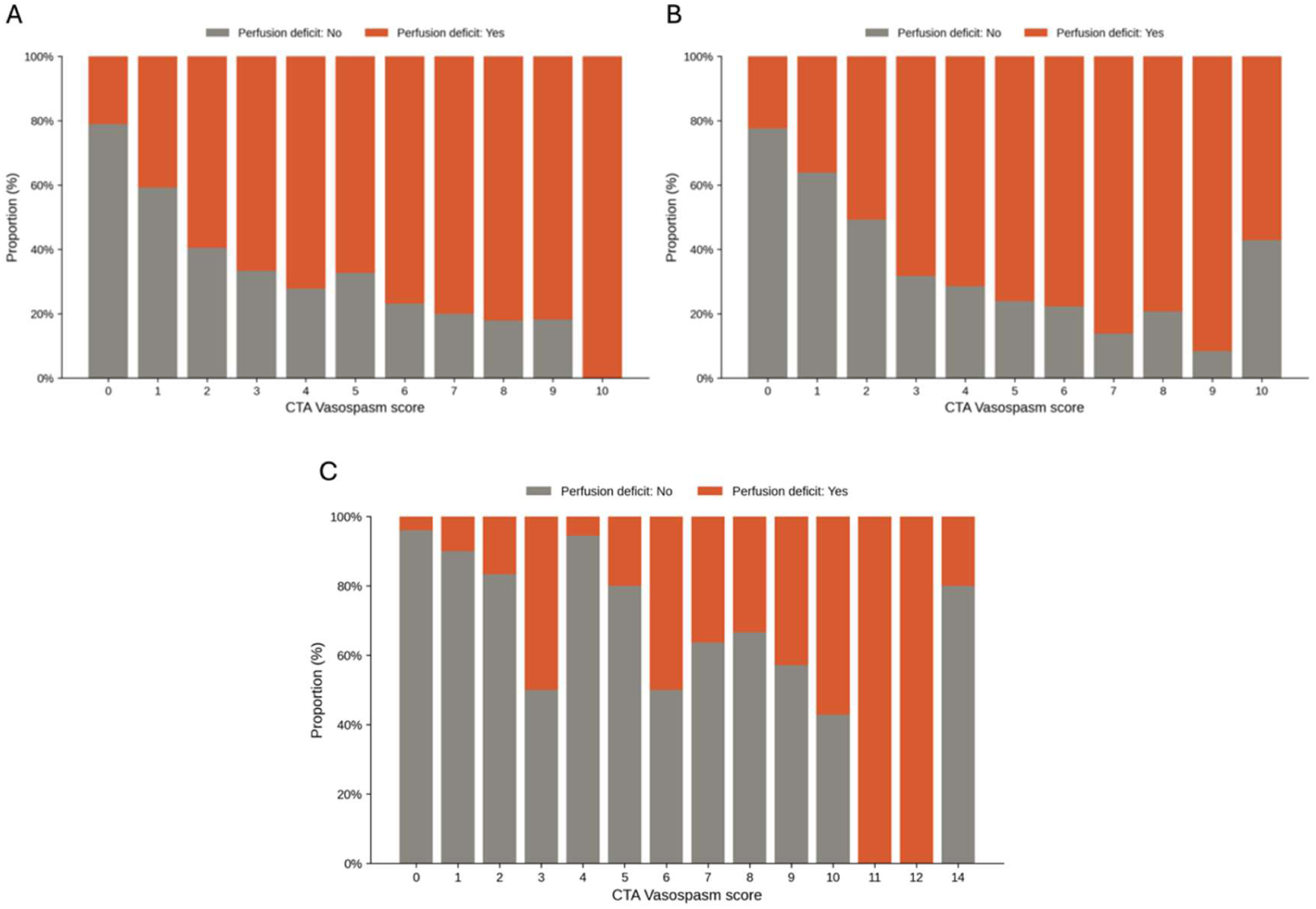
Proportion of examinations with a perfusion deficit in the left anterior circulation (A), right anterior circulation (B) and posterior circulation as a function of the CTA Vasospasm Score.

Using a GLMM, we found a significant association between increasing CTA Vaospasm Scores and the presence of a perfusion deficit in the corresponding territory (see Table 2). Higher odds ratios were observed between the CTA Vasospasm Score and perfusion deficits in the anterior circulation than in the posterior circulation. The Hunt&Hess grade, the number of days since hemorrhage and the modified Fisher scale were not associated with perfusions abnormalities. In a sensitivity analysis restricted to the 391 examinations performed between the third and 14th days after hemorrhage, the association between the CTA Vasospasm Score and perfusion deficits remained consistent with the primary analysis (left anterior circulation: OR [148], 95% CI [1.34 – 1.63]; right anterior circulation: OR [1.54], 95% CI [1.38 – 1.73]; posterior circulation: OR [1.31], 95% CI [1.17 – 1.45]).

**Table 1.** Distribution and severity of vasospasm.

| Vessel segment | Grade 1, n (%) | Grade 2, n (%) | Any vasospasm<br>(1+2), n (%) |
| --- | --- | --- | --- |
| ICA – left n = 503 | 90 (17.9%) | 49 (9.7%) | 139 (27.6%) |
| ICA – right n = 507 | 100 (19.7%) | 29 (5.7%) | 129 (25.4%) |
| M1 – left n = 501 | 147 (29.3%) | 134 (26.8%) | 281 (56.1%) |
| M1 – right n = 502 | 147 (29.3%) | 102 (20.3%) | 249 (49.6%) |
| M2 – left n = 511 | 127 (24.9%) | 110 (21.5%) | 237 (46.4%) |
| M2 – right n = 511 | 141 (27.6%) | 91 (17.8%) | 232 (45.4%) |
| A1 – left n = 495 | 138 (27.9%) | 138 (27.9%) | 176 (55.8%) |
| A1- right n = 484 | 130 (26.9%) | 135 (27.9%) | 265 (54.8%) |
| A2 – left n = 511 | 143 (28%) | 100 (19.6%) | 243 (47.6%) |
| A2 – right n = 511 | 155 (30.3%) | 94 (18.4%) | 249 (48.7%) |
| P1 – left n = 512 | 99 (19.3%) | 57 (11.2%) | 156 (30.5%) |
| P1 - right n = 508 | 83 (16.3%) | 64 (12.6%) | 147 (28.9%) |
| P2 - left n = 512 | 65 (12.7%) | 19 (3.7%) | 84 (16.4%) |
| P2 - right n = 512 | 65 (12.7%) | 19 (3.7%) | 84 (16.4%) |
| Basilar Artery n = 512 | 66 (12.9%) | 24 (4.7%) | 90 (17.6%) |
| V4 – left n = 507 | 36 (7.1%) | 11 (2.2%) | 47 (9.3%) |
| V4 – right n = 507 | 34 (6.7%) | 6 (1.2%) | 40 (7.9%) |

**Table 2.** Regression analysis showing the association between CTA Vasospasm Score and perfusion deficits in the left anterior circulation, right anterior circulation and posterior circulation, adjusted to Hunt&Hess grade, days since hemorrhage and modified Fisher score.

|  | Left anterior circulation |  | Right anterior circulation |  | Posterior circulation |  |
| --- | --- | --- | --- | --- | --- | --- |
|  | Odds ratio | 95%-CI | Odds ratio | 95%-CI | Odds ratio | 95%-CI |
| CTA Vasospasm Score | **1.49** | **1.36-1.63** | **1.51** | **1.38-1.67** | **1.37** | **1.24-1.53** |
| Hunt&Hess grade | 0.87 | 0.71-1.07 | 0.86 | 0.69-1.06 | 0.79 | 0.58-1.09 |
| Days since hemorrhage | 0.99 | 0.95-1.02 | 1 | 0.96-1.04 | 1 | 0.94-1.07 |
| Modified Fisher score | 1.00 | 0.78-1.3 | 0.99 | 0.75-1.3 | 1.30 | 0.84-2.00 |

Receiver operating characteristic analysis for predicting perfusion deficits revealed an area under the curve (AUC) of 0.77 (95%-CI 0.73-0.81) for the overall CTA Vasospasm Score. The optimal cut-off value, as determined by the Youden index, was 2.5, corresponding to a sensitivity of 84% and a specificity of 59%. Looking at the specific territories, we received an AUC of 0.77 (95%-CI 0.73-0.81) for the left anterior circulation (cut off: 1.5, sensitivity 82%, specificity 62%), 0.76 (95%-CI 0.72-0.8) for the right anterior circulation (cut off: 2.5, sensitivity 70%, specificity 74%), and 0.77 (95%-CI 0.70-0.84) for the posterior circulation (cut off: 2.5, sensitivity 74%, specificity 74%). Receiver operating characteristic (ROC) curves are shown in Figure 4.

**Figure 4.**
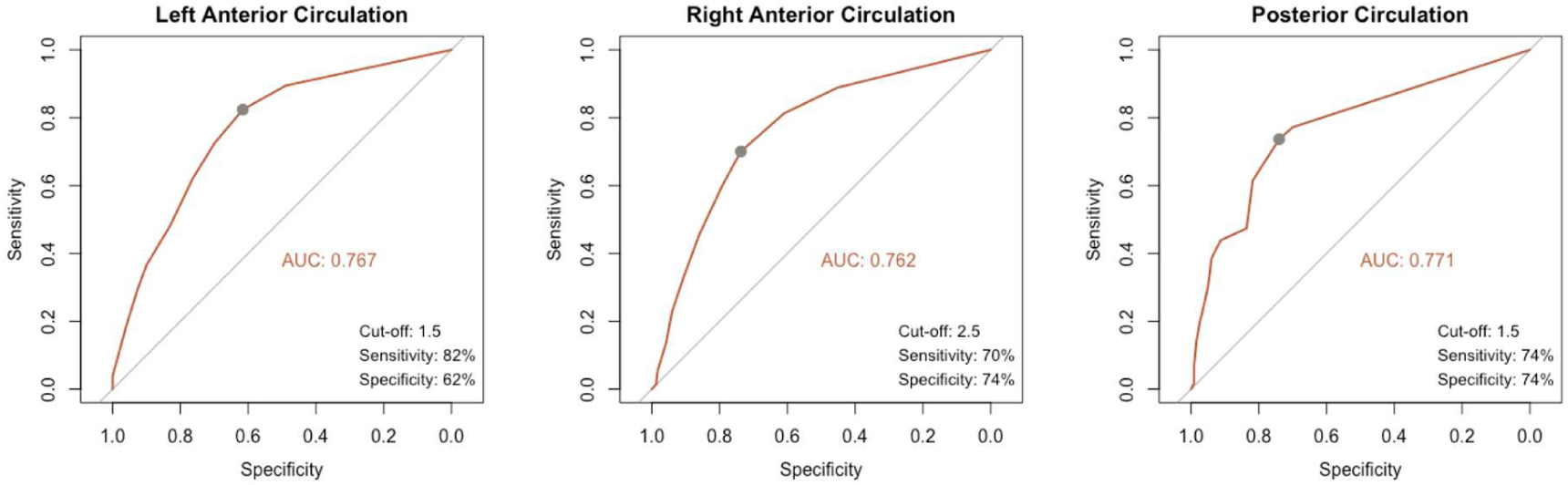
Receiver operating characteristic (ROC) curves for the CTA Vasospasm Score in predicting territorial perfusion deficits. Left anterior circulation, right anterior circulation, and posterior circulation. For each territory, the optimal cut-off value was determined using the Youden index, and the corresponding sensitivity and specificity are indicated. AUC = area under the curve.

## Discussion

CTA and CTP are commonly applied and recommended techniques to evaluate vasospasm and DCI in patients after aSAH.[3] Although the relationship between vasospasm and perfusion deficits is often assumed to be self-evident, our study offers a new perspective on the interplay between these two phenomena in patients with aSAH. The most significant findings from our results are: 1) the degree of cerebral vasospasm is associated with perfusion deficits within the territory of the spastic vessel, 2) the CTA vasospasm score offers a moderate to good sensitivity to predict perfusion deficits, and 3) there is a non-negligible proportion of examinations in which perfusion deficits can occur without vasospasm, and vice versa.

The finding that the degree of vascular narrowing is associated with perfusion deficits in the downstream vascular territory is not particularly surprising and has already been extensively studied in patients with focal stenoses and intracranial atherosclerotic disease.[9, 10] However, the difference in patients with vessel narrowing due to vasospasm is that vasospasm after aSAH is a diffuse and multifocal process and interpretation can be challenging.[11] One of the few studies which looked at the relationship between vasospasm grade and perfusion deficits was performed by Aralasmak et. al. Their results showed that there is a high likelihood (83%) of a perfusion deficit in the territory of vessel that demonstrates a vasospasm grade 2 (vessel narrowing > 50%). These results are in line with our results that show a likelihood of 81% of a perfusion deficit in patients with at least one vessel segment with vasospasm grade 2. The likelihood of a perfusion deficit decreased in patients with vasospasm grade 1. Here, slightly more than half of the patients (55%) showed a perfusion deficit.

In a more detailed approach to assess the relationship between the degree and pattern of vasospasm and perfusion abnormalities, we found an increasing proportion of patients with perfusion deficits with both increasing severity and multifocality of vasospasm. Mild focal vasospasm showed perfusion deficits in roughly one-third to two-fifths of examinations (35.8–40.4%), rising further in mild multifocal disease (54.3–58.8%), and reaching 71.6-74.4% in severe focal vasospasm before peaking at 81.2-82.2% in severe multifocal vasospasm. Taken together, these findings support a model in which the probability of perfusion deficit occurrence behaves as a continuum shaped jointly by two dimensions of vasospasm (severity and distribution), rather than being adequately captured by either parameter in isolation.

This has direct clinical implications: risk stratification and monitoring strategies for delayed cerebral ischemia should account for both the grade and the spatial pattern of vasospasm, and a purely severity-based classification may underestimate risk in patients with multifocal but individually mild-to-moderate spasm.

The CTA Vasospasm Score, introduced by van der Harst, incorporates both the severity and the spatial extent of vasospasm, combining these into a single composite score.[5] Van der Harst demonstrated an association between the CTA Vasospasm Score and the occurrence of DCI and unfavorable outcome in a relatively small study cohort. Our results align with those of van der Harst et al., as they demonstrate that the underlying cause, namely, the perfusion deficit and thus the microvascular hypoperfusion that can lead to neurological deficits and infarction [12], is directly associated with the CTA Vasospasm Score. Despite its clinical utility, the CTA Vasospasm Score demonstrated only moderate diagnostic accuracy in predicting territorial perfusion deficits, with sensitivity and specificity values in the ROC analysis falling short of the thresholds typically required for a stand-alone diagnostic criterion.

One of the key findings of our study is that a perfusion deficit was present in over 20% of patients even in the absence of a relevant detectable vasospasm. This finding carries pathophysiological and clinical implications since it supports the concept of DCI as a multifactorial process that extends beyond large-vessel vasospasm alone.[13] Microvascular dysfunction, including impaired cerebral autoregulation, microthrombosis, cortical spreading depolarizations, and neuroinflammatory cascades triggered by subarachnoid blood breakdown products, has increasingly been recognized as an independent contributor to perfusion compromise after aneurysmal SAH.[14–17] These mechanisms act at the level of the microcirculation and would not be captured by CTA, which is limited to depicting luminal narrowing of the visible, proximal and middle segments of the large vessels. Also, distal or segmental vasospasm affecting smaller, more peripheral arterial branches may fall below the resolution threshold of CTA or may simply not be adequately captured by a scoring system designed primarily around the major proximal vessel segments.

There are several limitations to our study. First, we used perfusion deficits as primary endpoints instead of occurrence of DCI or patients’ outcome, which is debatable regarding its clinical relevance. However, a perfusion deficit represents a direct pathophysiological correlate, whereas clinical deterioration or outcome can be influenced by a multitude of other factors. Also, instead of an ROI-based assessment of perfusion data, we used a visual, qualitative assessment of perfusion maps, which is a valid alternative and used by previous studies.[18] Second, the study is limited by its retrospective nature, including incomplete data and a potential selection bias. Imaging was not performed on predetermined days and main reason for performing the examination was predominantly a new clinical deficit, which can explain the high prevalence of perfusion deficits in our study. Finally, we also included vessels and territories from the posterior circulation, even though CTP evaluation may be limited in this area.

In conclusion, our results, on the one hand, clearly demonstrate the association between vasospasm and perfusion abnormalities, but on the other hand also indicate that microcirculatory abnormalities may have additional causes that are not captured by CTA. Therefore, we recommend acquiring a CTP in cases of clinical deterioration or in patients who cannot be clinically assessed, in order not to miss each patient’s ischemic risk.

## Conflict of Interest

FS, CH, BT, HSM, LD and VG have no relevant financial or non-financial interests to disclose. HK reports compensation from Eppdata, is a shareholder of Eppdata, and travel funding from Penumbra. MB, LM, and CT report compensation from Eppdata for consultant services. JF reports funding from the European Commission; personal consulting fees from Acandis, Cerenovus, Medtronic, Microvention, Phenox, Stryker, and Roche; consulting at Philips (no payments); payment or honoraria for lectures, presentations, speakers bureaus, manuscript writing or educational events from Penumbra and Tonbridge; support for attending meetings or travel from Medtronic and Penumbra; stock or stock options from Tegus Medical, Eppdata, and Vastrax; and participation in a Data Safety Monitoring Board or Advisory Board at Phenox (personal fees) and Stryker (personal fees) and is a past president of ESMINT.

## Funding

The authors state that this work has not received any funding.

## Data Availability

The data that support the findings of this study are not publicly available due to institutional and privacy restrictions but are available from the corresponding author upon reasonable request.

## Notes

### Competing Interest Statement

The authors have declared no competing interest.

### Clinical Trial

This study is a retrospective observational analysis of previously collected clinical and imaging data and does not report the results of a clinical trial. Therefore, prospective trial registration was not required.

### Author Declarations

Institutional Review Board of the Hamburg Chamber of Physicians (Ethikkommission der Ärztekammer Hamburg) (2022-300245-WF)

